# Effectiveness of fourth dose COVID-19 vaccine against the Omicron variant compared to no vaccination

**DOI:** 10.1101/2022.08.17.22278807

**Authors:** Jessie Zeng, Joshua Szanyi, Tony Blakely

**Affiliations:** Population Interventions Unit, Centre for Epidemiology and Biostatistics Research, Melbourne School of Population and Global Health, University of Melbourne, Parkville, Victoria, Australia

**Author notes:** Corresponding author: Tony Blakely, PhD, Population Interventions Unit, Centre for Epidemiology and Biostatistics Research, Melbourne School of Population and Global Health, University of Melbourne, 207-221 Bouverie St, Parkville, VIC 3010, Australia.

## Abstract

In response to enhanced immune evasion properties of the Omicron SARS-CoV-2 variant and waning COVID-19 vaccine effectiveness (VE), several jurisdictions have rolled out fourth dose vaccination programs. Using a system of logistic regression equations and VE estimates for a fourth dose relative to a third dose reported in an Israeli study, we estimated absolute vaccine effectiveness for third and fourth doses of mRNA COVID-19 vaccine (c.f. no vaccination) against Omicron, by clinical outcome. We found that a fourth dose restores or even enhances protection conferred by a third dose at the same time since vaccination.

## Introduction

Waning COVID-19 vaccine effectiveness (VE) and the capacity of the Omicron SARS-CoV-2 variant to evade pre-existing immunity have been major impediments to COVID-19 control efforts worldwide. In response, several countries have rolled out fourth dose COVID-19 vaccination programs^1-3^. Published studies have reported relative VE (rVE) against Omicron among individuals aged 60 years and older for a recent fourth dose of an mRNA-based vaccine, compared to a third dose administered at least four months prior^1-2^. One study thus far reported fourth dose VE relative to no vaccination^3^, which we term absolute VE (aVE). However, third and fourth dose aVE for comparable time periods are not available. We aimed to estimate the aVE of three- and four-dose COVID-19 vaccine regimens at the same time since vaccination.

## Methods

We previously published a logistic regression equation that predicts aVE, with waning over time, for a third dose of mRNA vaccine, regardless of primary course, against symptomatic infection and hospitalisation due to Omicron based on data published by the UK Health Security Agency (UKHSA) ^4^. We extended this equation to include age variation in VE, using a study reporting VE by age against the Delta variant^5^. We then incorporated the additional outcomes of any infection (asymptomatic and symptomatic) and mortality using data reported by the UKHSA^6^, creating a system of equations to estimate third dose VE against Omicron (BA.1) by disease severity, age, and time since vaccination.

We then applied these equations to estimate third dose aVE at three weeks post-vaccination and converted Magen (2022) et al’s reported fourth dose rVE into estimates of fourth dose aVE three weeks after vaccination, assuming an average interval of 20 weeks between third and fourth doses in the Magen et al study ^1-2^, using the formula:

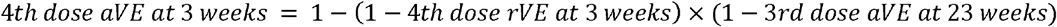

‘Expected’ aVE values and their uncertainty intervals were estimated using 10,000 iterations of Monte Carlo simulation propagating uncertainty in the reported fourth dose rVE and our logistic equation estimates of aVE for a third dose.

## Results

Estimated aVE against Omicron by clinical outcome at three weeks following the receipt of third and fourth doses is shown in *Figure*. As seen with a three-dose regimen, aVE following a fourth dose is higher with increasingly severe clinical outcomes. Estimated aVE following a fourth dose appears higher than after a third dose at the same time post-vaccination, but uncertainty intervals overlap.

**Figure.**
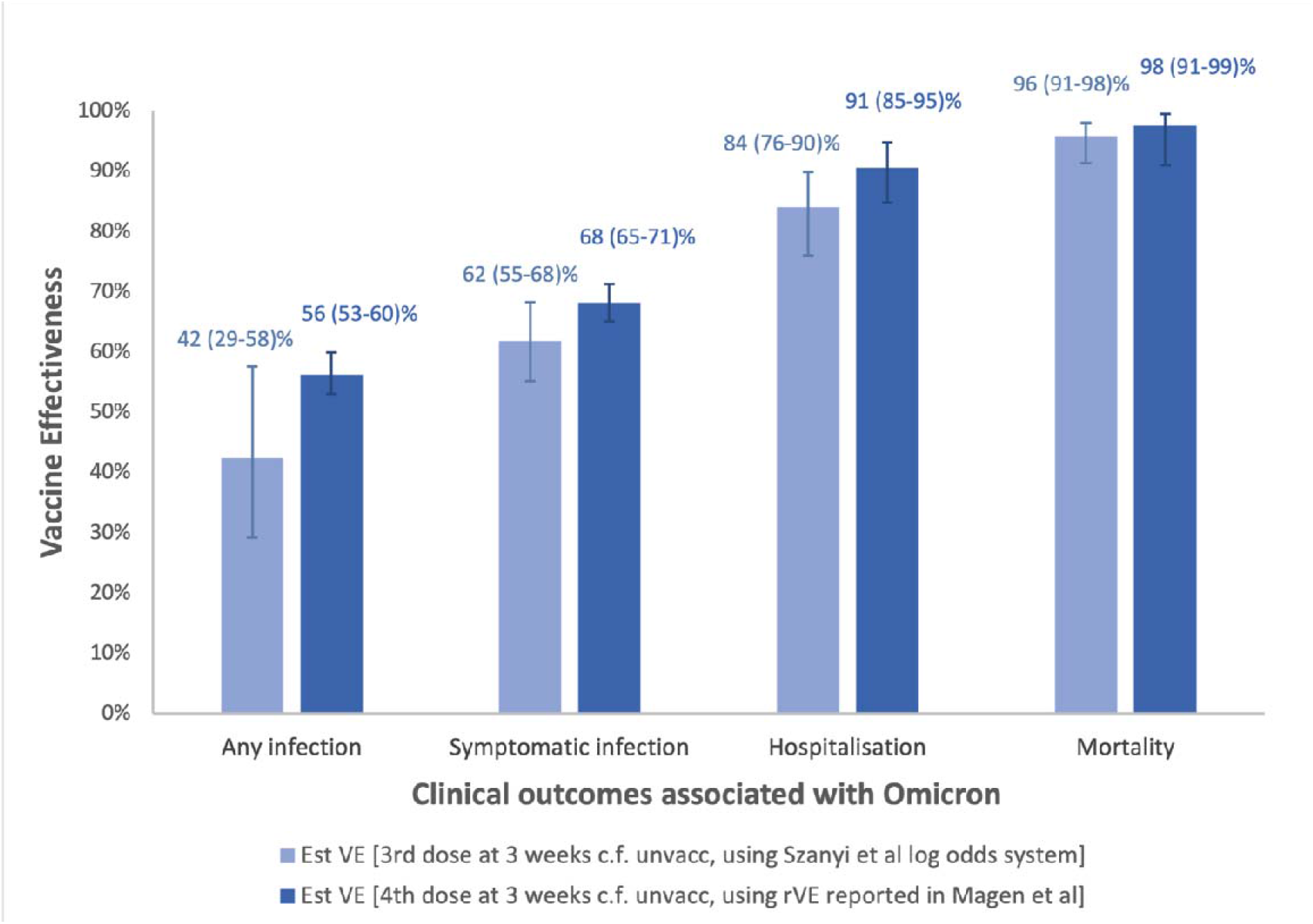
Vaccine effectiveness of Omicron-associated clinical outcomes at 3 weeks after third and fourth dose. Estimated aVE with 95% confidence intervals against clinical outcomes caused by the Omicron variant of SARS-CoV-2 3 weeks after a fourth vaccine dose (dark blue bars) compared to those 3 weeks after a third vaccine dose (light blue bars), with the unvaccinated as the reference category.

## Discussion

A fourth dose of an mRNA vaccine appears to restore protection conferred by a third dose at the same time since vaccination. The fourth dose boosting effect may even exceed the third dose for any infection and symptomatic infection, however, uncertainty intervals overlap. Our estimates are likely to be of interest to policy makers weighting up the benefits and costs of a fourth dose.

Our approach has limitations. Firstly, we imposed UKHSA data onto Israeli data, assuming a third dose in Israel has the same waning as a third dose in the UK. Nevertheless, these seem a reasonable assumption and it is difficult to conceive of an alternative method to estimate aVE, in the absence of any studies that directly compare individuals receiving third and fourth doses at the same point in time – each compared to those with no prior vaccination or infection. Secondly, our estimates concern VE against the BA.1 Omicron subvariant. Fourth dose aVE against newer subvariants may be reduced due to immune escape, although we expect that the patterns we estimate of restored or even somewhat higher aVE following a fourth dose likely hold.

## Data Availability

All data produced in the present study are available upon reasonable request to the authors

